## Supplemental Information for "Motivators and barriers to research participation for individuals with cerebral palsy and their families"

#### Supporting Information

##### S1 Table: Significant Pairwise Comparisons after Bonferroni Correction

| Variable | GMFCS Level α = 0.005 | CP Type α = 0.0083 | Responder Type α = 0.05 |
|---|---|---|---|
| ***Additional travel needs*** | | | |
| Time | | Di > Hemi<br>Quad > Hemi | |
| Breathing | Level V > Level IV<br>Level V > Level I | | |
| Transition | | | Adults > Parents |
| Seizure | Level V > Level IV<br>Level V > Level II | | Parents > Adults |
| Feeding | Level V > Level IV<br>Level V > Level III<br>Level V > Level II<br>Level V > Level I | Quad > Di<br>Quad > Hemi | Parents > Adults |
| Snacks | | | Parents > Adults |
| Medications | Level V > Level IV<br>Level V > Level III<br>Level V > Level II<br>Level V > Level I | | |
| Toileting | Level V > Level IV<br>Level V > Level III<br>Level V > Level II<br>Level V > Level I<br>Level IV > Level II | Quad > Di<br>Quad > Hemi | Parents > Adults |
| Transportation | Level V > Level II<br>Level V > Level I<br>Level IV > Level II<br>Level IV > Level I<br>Level III > Level II<br>Level III > Level I | Quad > Di<br>Quad > Hemi<br>Quad > Other<br>Di > Hemi | |
| Other | | | |
| ***Transportation*** | | | |
| Drive self | | Hemi > Di<br>Hemi > Quad | Parents > Adults |

| | | | |
|---|---|---|---|
| Family member drives | | | |
| Public transit | | | Adults > Parents |
| Ride service | | | Adults > Parents |
| Other | | | Adults > Parents |
| ***Locations*** | | | |
| Current clinic | | | Parents > Adults |
| New clinic | | Hemi > Quad | |
| Park | Level I > Level II<br>Level I > Level III<br>Level I > Level IV<br>Level I > Level V | | Parents > Adults |
| Lab | | Hemi > Quad | |
| School | | | Parents > Adults |
| Home | | | |
| Other | | | |

***Abbreviations:** GMFCS = Gross Motor Function Classification System, di = diplegia, hemi = hemiplegia, quad = quadriplegia

### Participation in Research Survey

Thank you for your willingness to answer questions about motivators and barriers to participation in cerebral palsy (CP) research. Please complete the survey below. If you are a parent of an individual with CP, we encourage you to discuss the survey questions with your child and fill out the questions together.

If you have more than one child with CP, please choose one child to answer the demongraphics section about, and then consider the whole family in answering the questions that follow.

Finally, please try to answer these questions without considerations for the current COVID-19 crisis unless otherwise specified.

Thank you for your contributions to better research study design!

#### Demographic information

Do you live in the United States?

◯ Yes
◯ No

What is your zip code?

__________________________________

Please select the most appropriate description below.

◯ I am an adult with cerebral palsy.
◯ I am the parent or guardian of a minor with cerebral palsy.

Do you consider yourself to be Hispanic or Latino, defined as a person of Cuban, Mexican, Puerto Rican, South or Central American, or other Spanish culture or

origin, regardless of race? Select one.
(for more information, see https://grants.nih.gov/grants/guide/notice-files/NOT-OD-20-031.html and https://www.govinfo.gov/content/pkg/FR-1997-10-30/html/97-28653.htm)

◯ Hispanic or Latino
◯ Not Hispanic or Latino
◯ Do not wish to report

Do you consider yourself to be Hispanic or Latino, defined as a person of Cuban, Mexican, Puerto Rican, South or Central American, or other Spanish culture or

origin, regardless of race? Select one or more.
(for more information, see https://grants.nih.gov/grants/guide/notice-files/NOT-OD-20-031.html and https://www.govinfo.gov/content/pkg/FR-1997-10-30/html/97-28653.htm)

☐ American Indian or Alaska Native. A person having origins in any of the original peoples of North, Central, or South America, and who maintains tribal affiliation or community attachment.
☐ Asian. A person having origins in any of the original peoples of the Far East, Southeast Asia, or the Indian subcontinent, including, for example, Cambodia, China, India, Japan, Korea, Malaysia, Pakistan, the Philippine Islands, Thailand, and Vietnam.
☐ Black or African American. A person having origins in any of the black racial groups of Africa. Terms such as "Haitian" or "Negro" can be used in addition to "Black" or "African American."
☐ Native Hawaiian or Other Pacific Islander. A person having origins in any of the original peoples of Hawaii, Guam, Samoa, or other Pacific Islands.
☐ White. A person having origins in any of the original peoples of Europe, the Middle East, or North Africa.
☐ Do not wish to report

Please select the appropriate cerebral palsy diagnosis.

◯ hemiplegia
◯ diplegia
◯ quadriplegia
◯ other

Please state the name of the cerebral palsy diagnosis

__________________________________

Please select the current age of the person with cerebral palsy.

◯ 0-1 years
◯ 2-3 years
◯ 4-5 years
◯ 6-11 years
◯ 12-17 years
◯ 18-30 years
◯ 31-50 years
◯ 51+ years

Please select the most appropriate GMFCS level (Gross Motor Functional Classification System) description. If you are unfamiliar with these terms, you can also go to this website link for more information.

◯ Level I - Can sit on own, moves by walking without a walking aid; is able to balance in sitting when using both hands to play; can move in and out of sitting and standing positions without help from an adult; and prefers to move by walking .
◯ Level II - Can sit on own, usually moves by walking with a walking aid; may have difficulty with sitting balance when using both hands to play; can get in and out of sitting positions on own; can pull to stand and cruise holding onto furniture; and can crawl, but prefers to move by walking.
◯ Level III - Can sit on own, walk short distances with a walking aid (such as a walker, rollator, crutches,canes, etc.); may need help from an adult for steering and turning when walking with an aid; usually sits on floor in a "W-sitting" position; may need help from an adult to get into sitting; may pull to stand and cruise short distances, and prefers to move by creeping and crawling.
◯ Level IV - Can sit on own when placed on the floor; can move within a room; uses hands for support to maintain sitting balance; usually uses adaptive equipment for sitting and standing; and moves by rolling, creeping on stomach or crawling.
◯ Level V - Has difficulty controlling head and trunk posture in most positions; uses specially adapted seating to sit comfortably; and has to be lifted by another person to move about .

Please select the most appropriate GMFCS level (Gross Motor Functional Classification System) description. If you are unfamiliar with these terms, you can also go to this website link for more information.

◯ Level I - Can walk on their own without using a walking aid, including fairly long distances, outdoors and on uneven surfaces; can move from the floor or a chair to standing without using their hands for support; can go up and down stairs without needing to hold the handrail; is beginning to run and jump.
◯ Level II - Can walk on their own without using a walking aid, but has difficulty walking long distances or on uneven surfaces; can sit in a normal adult chair and use both hands freely; can move from the floor to standing without adult assistance; needs to hold the handrail when going up or down stairs; is not yet able to run and jump.
◯ Level III - Can walk on their own using a walking aid (such as a walker, rollator, crutches, canes, etc.); can usually get in and out of a chair without adult assistance; may use a wheelchair when traveling long distances or outside; finds it difficult to climb stairs or walk on an uneven surface without considerable help.
◯ Level IV - Can sit on their own but does not stand or walk without significant support and adult supervision; may need extra body / trunk support to improve arm and hand function; usually needs adult assistance to get in and out of a chair; may achieve self-mobility using a powered wheelchair or is transported in the community.
◯ Level V - Has difficulty sitting on their own and controlling their head and body posture in most positions; has difficulty achieving any voluntary control of movement; needs a specially-adapted supportive chair to sit comfortably; has to be lifted or hoisted by another person to move.

Please select the most appropriate GMFCS level (Gross Motor Functional Classification System) description. If you are unfamiliar with these terms, you can also go to  this website link  for more information.

- ◯ Level I - Can walk on their own without using walking aids; can go up or down stairs without needing to hold the handrail and walks wherever they want to go (including uneven surfaces, slopes or in crowds); can run and jump although their speed, balance, and coordination may be slightly limited.
- ◯ Level II - Can walk on their own without using walking aids; needs to hold the handrail when going up or down stairs and often finds it difficult to walk on uneven surfaces, slopes or in crowds.
- ◯ Level III - Can stand on their own and only walks using a walking aid (such as a walker, rollator, crutches, canes, etc.); finds it difficult to climb stairs, or walk on uneven surfaces; may use a wheelchair when traveling for long distances or in crowds.
- ◯ Level IV - Can sit on their own; does not stand or walk without significant support and therefore relies mostly on wheelchair at home, school and in the community; often needs extra body / trunk support to improve arm and hand function; may achieve self-mobility using a powered wheelchair.
- ◯ Level V - Has difficulty sitting on their own and controlling their head and body posture inmost positions; has difficulty achieving any voluntary control of movement; needs a specially supportive chair to sit comfortably; has to be lifted or hoisted by another person to move.

---

Please select the most appropriate GMFCS level (Gross Motor Functional Classification System) description. If you are unfamiliar with these terms, you can also go to  this website link  for more information.

- ◯ Level I - Can walk on their own without using walking aids; can go up or down stairs without needing to hold the handrail and walks wherever they want to go (including uneven surfaces, slopes or in crowds); can run and jump although their speed, balance, and coordination may be limited.
- ◯ Level II - Can walk on their own without using walking aids therefore walks in most settings; needs to hold the handrail when going up or down stairs and often finds it difficult to walk on uneven surfaces, slopes or in crowds; may occasionally prefer to use a walking aid (such as a cane or crutch) or a wheelchair to travel quickly or over longer distances.
- ◯ Level III - Can stand on their own and only walks using a walking aid(such as a walker, rollator, crutches, canes, etc.); finds it difficult to climb stairs, or walk on uneven surfaces without support; uses a variety of means to move around depending on the circumstances and prefers to use a wheelchair to travel quickly or over longer distances.
- ◯ Level IV - Can sit with some pelvic and trunk support; does not stand or walk without significant support and therefore always relies on wheelchair when outdoors; can achieve self-mobility using a powered wheelchair; can crawl or roll to a limited extent to move around indoors.
- ◯ Level V - Has difficulty sitting on their own and controlling their head and body posture in most positions; has difficulty achieving any voluntary control of movement; needs a specially adapted chair to sit comfortably and be transported anywhere; has to be lifted or hoisted by another person or special equipment to move.

---

Please select the most appropriate GMFCS level (Gross Motor Functional Classification System) description. If you are unfamiliar with these terms, you can also go to  this website link  for more information.

- ◯ Level I - Can walk on my own without using walking aids; can go up or down stairs without needing to hold the handrail and walk wherever I want to go (including uneven surfaces, slopes or in crowds); can run and jump although my speed, balance, and coordination may be limited.
- ◯ Level II - Can walk on my own without using walking aids and therefore walk in most settings; need to hold the handrail when going up or down stairs; often find it difficult to walk on uneven surfaces, slopes or in crowds and may occasionally prefer to use a walking aid (such as a cane or crutch) or a wheelchair to travel quickly or over longer distances.
- ◯ Level III - Can stand on my own and only walk using a walking aid(such as a walker, rollator, crutches, canes, etc.); find it difficult to climb stairs, or walk on uneven surfaces without support; use a variety of means to move around depending on the circumstances and prefer to use a wheelchair to travel quickly or over longer distances.
- ◯ Level IV - Can sit on my own; do not stand or walk without significant support and therefore always rely on wheelchair when outdoors; can achieve self-mobility using a powered wheelchair; can crawl or roll to a limited extent to move around indoors.
- ◯ Level V - Have difficulty sitting on my own and controlling my head and body posture in most positions; have difficulty achieving any voluntary control of movement; need a specially adapted chair to sit comfortably and be transported anywhere; have to be lifted or hoisted by another person or special equipment to move.

Please select the sex of the person with cerebral palsy.

◯ male
◯ female
◯ do not wish to report

#### Research Interest

How important do you think research is in the care of individuals with CP?

not important at all | very important

(Place a mark on the scale above)

How much do you value your or your child's ability to participate in research studies pertaining to CP?

not at all | very much

(Place a mark on the scale above)

Below are some examples of different types of cerebral palsy research studies.
If you were to receive a study flyer or email about the following types of studies, which ones would you be likely to contact researchers to learn more about? Please select all that apply.

☐ Activity monitoring during your daily activities (with devices such as FitBits, AppleWatches, etc)
☐ Robotic games to measure movement
☐ Study of the effect of a drug or medication, where you would definitely receive the medication
☐ Study of a new treatment, where you may or may not receive the new treatment
☐ Study of a physical or occupational therapy treatment, where you would definitely get the treatment
☐ Study of speech therapy treatment, where you would definitely get the treatment
☐ Survey, focus group, or other online study
☐ Technology that take pictures of the muscle or bone (example: ultrasound or magnetic resonance imaging - MRI)
☐ Technology that take pictures of the brain (example: magnetic resonance imaging - MRI)
☐ Other

Please explain what other study would be of interest to you:

__________________________________

Research studies can focus on different areas or functions of the body.

Which area of research focus would you be most interested in participating in, or hearing more about? Please select all that apply.

☐ Arms or hands
☐ Brain or nerves
☐ Continence or toileting
☐ Digestion or nutrition
☐ Genetics or stem cells
☐ Heart or lungs
☐ Language or learning
☐ Legs or feet
☐ Movement or fitness
☐ Muscles
☐ Pain
☐ Other

Please explain what other study focus would be of interest to you:

__________________________________

**Practical considerations and accessibility**

Which locations/areas would you be most likely to agree to participate in a study? Consider where you would be most comfortable for reasons that are most relevant to you. Select all that apply.

☐ Clinic/hospital where you are already receiving services
☐ Clinic/hospital where you are not receiving services
☐ Park or community center
☐ Research lab at a University
☐ School
☐ Your home
☐ Other

---

What other location would be desirable for you?

__________________________________

---

Are there any locations you would definitely not consider going to? Select all that apply.

☐ Clinic/hospital where you are already receiving services
☐ Clinic/hospital where you are not receiving services
☐ Park or community center
☐ Research lab at a University
☐ School
☐ Your home

---

How do you typically get yourself or your child to/from medical appointments? Check all that apply.

☐ Drive Self
☐ Family member drives
☐ Public transportation (bus, metro/subway, high speed rail)
☐ Uber/Lyft/Taxi
☐ Other

---

What other transportation do you use?

__________________________________

---

How long does it typically take you to get to a medical appointment?

◯ less than 30 minutes
◯ 30 minutes to 1 hour
◯ 1 to 1.5 hours
◯ 1.5 to 2 hours
◯ More than 2 hours

Which of the following are things that you need to pack or consider when leaving your home? Select all that apply.

- ☐ additional time needed to direct task of leaving the house
- ☐ breathing items such as tracheostomy care, inhalers, etc
- ☐ concerns about ability to adjust to a new environment or transition between environments
- ☐ concerns or items related to potential seizure activity
- ☐ feeding items for g-tube
- ☐ feeding items such as snacks, drinks, etc
- ☐ medications
- ☐ toileting items such as diapers, catheter supplies, etc
- ☐ transportation items such as a wheelchair or stroller
- ☐ other

---

Please specify other considerations.

__________________________________

---

Would you be willing to make an overnight trip or an extended trip to participate in a research study?

- ○ Yes
- ○ No
- ○ Maybe

---

Because you selected that you may be willing to travel, what does this depend on?

__________________________________

---

Is your willingness to travel to participate in a research study dependent on the type of study (whether it has the potential to benefit the person with CP or has a purpose to generally understand more about CP)?

- ○ Yes
- ○ No

---

For a study that has the potential to offer a direct benefit to the participant, what is the maximum time you would spend to travel from your home to the study?

- ○ 30 min
- ○ 1 hour
- ○ 1.5 hours
- ○ 2 hours
- ○ More than 2 hours

---

For a study that is seeking to understand more about CP, what is the maximum time you would spend to travel from your home to the study?

- ○ 30 min
- ○ 1 hour
- ○ 1.5 hours
- ○ 2 hours
- ○ More than 2 hours

What is the maximum time you would spend to travel from your home to the study?

◯ 30 min
◯ 1 hour
◯ 1.5 hours
◯ 2 hours
◯ More than 2 hours

In general, I think it's important for research participants to be compensated or paid for their time.

not important at all — very important

*(Place a mark on the scale above)*

What do you feel is appropriate compensation for a family's time for participating in research?

Enter a value in whole dollar amounts per hour (for example enter "10" for $10/hour).

__________________________________

How important is it to you that the cost of travel for the research study be paid for or reimbursed if the study were local?

would not affect my decision — key factor in my decision

*(Place a mark on the scale above)*

How important is it to you that the cost of travel for the research study be paid for or reimbursed if the study required a longer distance trip?

would not affect my decision — key factor in my decision

*(Place a mark on the scale above)*

Does your willingness to participate depend on time of year? Please check all times of that you would consider participation in research.

☐ Weekends during the school year
☐ Weekdays during the school year
☐ Summer break
☐ Spring break
☐ Winter break
☐ Other school non-attendance days

Are there other family members (children or others) who would need care arranged for in order for the person with CP to participate in a research study?

◯ Yes
◯ No

Are there people who would need to take time off of work (parent, participant, others) in order for the individual with CP to participate in a research study?

◯ Yes
◯ No

What amount of time in one day would be reasonable to participate in a study for you/your child? Consider the time spent in a laboratory or clinic, you/your child's attention span, etc. Check all that seem reasonable:

☐ 30 min
☐ 1 hour
☐ 2 hours
☐ 3 hours
☐ 4 hours
☐ 5 hours
☐ 6 hours
☐ 7 hours
☐ 8 hours

What is the maximum number of visits you would be willing to participate in for a single research study?

○ 1 visit
○ 2 visits
○ 3 visits
○ 4 visits
○ 5+ visits

Would you consider participating in a study that requires repeated visits over a number of months or years?

○ Yes
○ No

**Which of the following treatments have you or has your child received? Select all relevant responses from this table:**

| | arms | legs | spine or trunk |
|---|---|---|---|
| Bony surgery (examples include derotation osteotomies, bone lengthening) | ☐ | ☐ | ☐ |
| Soft tissue surgery (examples include muscle lengthening, tendon transfers) | ☐ | ☐ | ☐ |
| Neural surgery (examples include selective dorsal rhizotomy) | ☐ | ☐ | ☐ |
| Botox or other injections within the last year | ☐ | ☐ | ☐ |
| Spasticity medication not received through injections (examples include Baclofen) currently | ☐ | ☐ | ☐ |
| Physical or Occupational Therapy currently | ☐ | ☐ | ☐ |
| Intensive therapy programs or camps (examples include constraint induced movement therapy (CIMT), hand arm bimanual therapy (HABIT), gait camp) | ☐ | ☐ | ☐ |

**Past research experience**

Have you participated in research studies in the past?

〇 Yes
〇 No

Briefly, can you describe studies you have participated in?

Have your previous studies included ones conducted in the Department of Physical Therapy and Human Movement Sciences (NUPTHMS) at Northwestern University? The address of this location is 645 North Michigan Avenue Chicago, IL 60611.

〇 Yes
〇 No

What motivated your choice to participate in studies at NUPTHMS?

How would you rate your experience participating in research at NUPTHMS?

mostly negative — largely positive

*(Place a mark on the scale above)*

How easy was it to get to NUPTHMS?

extremely difficult — very easy

*(Place a mark on the scale above)*

How was the communication between you and the researcher at NUPTHMS?

very poor — excellent

*(Place a mark on the scale above)*

How was your comfort level with the facilities at NUPTHMS?

very poor — excellent

*(Place a mark on the scale above)*

I felt that participating in research at NUPTHMS changed my perspective on the necessity/efficacy of research for cerebral palsy.

completely disagree — completely agree

*(Place a mark on the scale above)*

What could have improved your experience participating in research at NUPTHMS?

In general, what is the biggest motivator for you or your child to participate in research?

In general, what is the biggest barrier for you or your child to participate in research?

What would you hope to get out of your/your child's participation in research?

**You were asked to fill out this survey without thinking about the current COVID-19 pandemic and associated restrictions to typical life. However, the pandemic has changed much about attitudes towards social interactions. Assuming the following activities are locally allowed at the time, when do you think you might feel comfortable resuming these activities for you or your child?**

| | now | lifting of stay at home order | following availability of widespread testing | following vaccinat on availability | I would not be willing in the foreseeable future |
|---|---|---|---|---|---|
| Medical appointment for new concern | ☐ | ☐ | ☐ | ☐ | ☐ |
| Personal care appointment (eg. hair, nails, grooming) | ☐ | ☐ | ☐ | ☐ | ☐ |
| Recreational or social activity (eg. movie theater, playdate) | ☐ | ☐ | ☐ | ☐ | ☐ |
| Research study participation with potential direct benefit | ☐ | ☐ | ☐ | ☐ | ☐ |
| Research study participation without direct benefit | ☐ | ☐ | ☐ | ☐ | ☐ |
| Routine medical appointment (eg. check up) | ☐ | ☐ | ☐ | ☐ | ☐ |

Has your experience with COVID-19 impacted how you would feel about participating in research within the next 6 months?
